# Nicotine strength of e-liquids used by adult vapers in Great Britain: a population survey 2016 to 2024

**DOI:** 10.1101/2024.03.13.24304222

**Authors:** Sarah E. Jackson, Jamie Brown, Lion Shahab, Deborah Arnott, Linda Bauld, Sharon Cox

## Abstract

**Background/Aims:** In March-2024, the UK government announced plans to introduce a new Vaping Products Duty that will tax e-liquids based on their nicotine strength. This study examined trends in the nicotine strength of e-liquids used by adult vapers and differences in those currently used across relevant subgroups.

**Design:** Nationally-representative, cross-sectional household survey, July-2016 to January-2024.

**Setting:** Great Britain.

**Participants:** 7,957 adult vapers.

**Main outcome measures:** Participants were asked whether the e-cigarette they mainly use contains nicotine and the e-liquid strength. We used logistic regression to estimate time trends in different nicotine strengths used (no nicotine/>0-≤6/7-11/12-19/≥20 mg/ml), overall in England and stratified by main device type (disposable/refillable/pod), age (≥18y), and smoking status. We explored current differences in nicotine strength among those surveyed between January-2022 and January-2024 in Great Britain by main device type, age (≥16y), gender, occupational social grade, history of ≥1 mental health conditions, smoking status, and (among past-year smokers) level of cigarette addiction.

**Results:** The proportion of vapers in England using high-strength (≥20mg/ml) e-liquids increased from an average of 3.8% [95%CI 2.9-5.0%] up to June-2021 to 32.5% [27.9-37.4%] in January-2024 (when 93.3% reported using exactly 20mg/ml). This rise was most pronounced among those using disposable e-cigarettes, those aged 18-24y, and all smoking statuses (including never smokers) except long-term (≥1y) ex-smokers. Of those surveyed in 2022-24 in Great Britain, overall, 89.5% [88.1-90.8%] said they usually used e-cigarettes containing nicotine, 8.7% [7.5-10.0%] used nicotine-free e-cigarettes, and 1.8% [1.2-2.4%] were unsure. The proportion using ≥20mg/ml was higher among those mainly using disposable (47.9%) compared with pod (16.3%) or refillable (11.5%) devices; never smokers (36.0%), current smokers (28.8%), or recent (<1y) ex-smokers (27.4%), compared with long-term ex-smokers (13.9%); and younger (16-24y; 44.2%) compared with older (≥25y; range 9.4-25.1%) age groups. There were no notable differences across other subgroups of interest.

**Conclusions:** Use of high-strength nicotine e-liquids in England has increased sharply in recent years. Most adult vapers in Great Britain use e-cigarettes that contain nicotine but different subgroups use different strengths: they tend to be higher among those who mainly use disposable devices, those aged 16-24y, and lower among long-term ex-smokers.

## Introduction

In Great Britain, the prevalence of e-cigarette use (‘vaping’) has risen rapidly among adolescents and young adults since 2021.^1–3^ This has largely been attributed to the introduction of new disposable e-cigarettes (vapes) to the market.^2^ These products are easy to use, have colourful design and branding, come in a variety of flavours, and typically contain high levels of nicotine, delivered in a palatable nicotine-salts e-liquid.^4–6^ They are also cheaper to buy than both cigarettes and refillable e-cigarettes – one of the most popular brands with underage vapers, Elf Bar 600, can be found online for £2.99 (US$3.80, €3.50) and in convenience stores and supermarkets for £5.99 (US$7.70, €7.00) – making them more affordable for experimental use.

The UK government has made reducing youth vaping a key public health policy priority.^7^ In January 2024, the Prime Minister announced a ban on disposable e-cigarettes as part of a package of measures designed to tackle the rise in youth vaping.^8,9^ In March 2024, the Chancellor of the Exchequer announced in the Spring budget that a new Vaping Products Duty would also be introduced from October 2026 in an effort to make vaping less affordable to children.^10^ According to the consultation document, which sets out the proposal for how the duty will be designed and implemented, the Vaping Products Duty will be an excise duty levied on the e-liquid in e-cigarettes with higher levels of duty applied to higher-strength nicotine e-liquids (proposed to be £1 per 10ml for nicotine-free e-liquids, £2 per 10ml for e-liquids that contain up to 10.9 mg of nicotine per ml, and £3 per 10ml for e-liquids that contain 11 mg or more nicotine per ml).^10^ The rationale for the differential tax rate is that ‘given the known harms caused by nicotine addiction, the government’s intention is also to encourage consumers to reduce their nicotine intake by switching to lower or nicotine-free options, further supporting health objectives’.^10^ Some evidence suggests that use of products with higher nicotine strengths is associated with greater symptoms of dependence (e.g., frequency of vaping, urges to vape, and perceived vaping addiction) among young vapers.^11^

Ministers have acknowledged the need to try and strike the right balance with a price increase that acts as a deterrent but ensures vaping remains a more affordable option than smoking to encourage adult smokers to switch to the less harmful product.^9^ To that end, the UK government concurrently announced a one-off increase on the duty on tobacco (by £2 per 100 cigarettes or 50 grams of tobacco) in October 2026 to coincide with the introduction of the Vaping Products Duty.^10^ However, even if they remain less expensive than tobacco, taxing higher-strength nicotine e-liquids at higher rates could have unintended consequences for people who smoke and those who have switched from smoking to vaping. Higher-strength e-liquids provide better relief from withdrawal and satisfy cravings for tobacco^12^ and may therefore be more effective for helping prevent relapse.^13^ Making these products more expensive could disincentivise their use and drive vapers towards cheaper, lower-strength e-liquids. This could potentially undermine smoking cessation^14^ or result in increased use of e-liquid to compensate^15^ (including among young vapers not trying to quit smoking), thus increasing potential toxicant exposure and associated risks to health.^16^ It could also prompt them to source illicit higher strength products and potentially also to mix their own e-liquids, which poses potential safety risks.^17^ These responses may be more likely among vapers from disadvantaged groups (e.g., those working in lower paid jobs or with mental health conditions), who tend to be more dependent on nicotine^18,19^ and have lower disposable incomes.

Understanding what nicotine strengths adult vapers in Great Britain are currently using, prior to the introduction of the Vaping Products Duty, and how this differs across subgroups of vapers, can offer insight into who will be most affected by the duty. In line with the European Union Tobacco Products Directive (TPD), the maximum nicotine concentration permitted in e-liquids for sale as consumer products in Great Britain is 20 mg/ml.^16^ A representative survey of adults in England in 2021 indicated the most popular strength of e-liquid was ≤6 ml/ml (used by 39.9% of vapers), with just 5.4% using 20 mg/ml or more,^16^ but it is likely that this may have changed since high-strength disposable e-cigarettes have become popular. This study aimed to:

1. Estimate trends in the nicotine strength of e-liquids used among adult vapers in England between 2016 and 2023, overall and by the main device type used and smoking status.
2. Provide up-to-date descriptive information on the use of different e-cigarette nicotine strengths among adult vapers in Great Britain in 2022-24, overall and by the main device type used, age, gender, socioeconomic position, history of mental health conditions, smoking status, and (among past-year smokers) level of cigarette dependence.
3. Understand which groups would be most affected by the proposed Vaping Products Duty structure.

## Methods

### Design

Data were drawn from the Smoking Toolkit Study.^20,21^ This is a repeat cross-sectional survey of adults (≥16 years) that captures a broad range of data on smoking and vaping. It began in England in 2006 (*n*~1,700 per month) and expanded to cover Wales (*n*~300) and Scotland (*n*~450) from October 2020. Each month, a new sample is recruited using a hybrid of random probability and quota sampling. Comparisons with other national surveys and sales data indicate that key variables such as sociodemographic characteristics, smoking prevalence, and cigarette consumption are nationally representative.^20,22^

Interviews were conducted face-to-face up to the start of the Covid-19 pandemic. Social distancing restrictions meant no data were collected in March 2020 and data collection pivoted to telephone interviews from April 2020 onwards. The two data collection modalities show good comparability: when social distancing restrictions were lifted, we ran a parallel telephone and face-to-face survey wave and yielded similar estimates for key sociodemographic, smoking, and nicotine product use measures.^23^ Data were not collected from 16- and 17-year-olds between April 2020 and December 2021.

For the present analyses, we selected data from two samples of participants who reported current vaping at the time of the survey. The trend analyses focused on respondents in the period from July 2016 (the first wave to assess nicotine strength) to January 2024 (the most recent data at the time of analysis). We restricted this sample to those living in England and aged ≥18 for consistency across the time series, given that the Wales and Scotland data collection began in 2020 and 16- and 17-year-olds were not included in every wave. The analyses of current nicotine strengths used in 2022-24 focused on all respondents living in Great Britain and aged ≥16 in the period from January 2022 to January 2024 (the most recent two years of data available, after new disposable e-cigarettes became popular^2^).

Nicotine strength was not assessed in certain waves (May, June, August, September, November, and December 2022 and February, March, May, August, September, November, and December 2023), so we excluded participants surveyed in these waves from our analytic samples.

### Measures

Vaping status was assessed within several questions asking about use of a range of nicotine products. Current smokers were asked ‘Do you regularly use any of the following in situations when you are not allowed to smoke?’; current smokers and those who have quit in the past year were asked ‘Can I check, are you using any of the following either to help you stop smoking, to help you cut down or for any other reason at all?’; and non-smokers were asked ‘Can I check, are you using any of the following?’. Those who reported using an e-cigarette in response to any of these questions were considered current vapers and formed our analytic sample.

Nicotine strength was assessed with two questions. The first asked: ‘Does the electronic cigarette or vaping device you mainly use contain nicotine?’ with response options ‘yes’, ‘no’, and ‘don’t know’. Those who responded yes to this question were then asked: ‘What strength is the e-liquid that you mainly use in your electronic cigarette or vaping device?’ with response options ‘6 mg/ml (0.6%) or less’, ‘7 mg/ml (0.7%) to 11 mg/ml (1.1%)’, ‘12 mg/ml (1.2%) to 19 mg/ml (1.9%)’, ‘20 mg/ml (2.0%) or more’, and ‘don’t know’. From the most recent survey (January 2024), the response option ‘20 mg/ml (2.0%) or more’ has been replaced with ‘20 mg/ml (2.0%)’ and ‘more than 20 mg/ml (2.0%)’ to distinguish between those using e-liquids with nicotine strengths at vs. exceeding the legal limit. The vast majority (*n*=56/60, 93.3%) of participants surveyed in this wave who reported using 20 mg/ml or more said they used 20 mg/ml exactly (i.e., the maximum legal limit).

Device type was assessed with the question: ‘Which of the following do you mainly use…?’ Response options were:

- Refillable – ‘An e-cigarette or vaping device with a tank that you refill with liquids (rechargeable)’ or ‘A modular system that you refill with liquids (you use your own combination of separate devices: batteries, atomizers, etc.)’
- Disposable – ‘A disposable e-cigarette or vaping device (non-rechargeable)’
- Pod – ‘An e-cigarette or vaping device that uses replaceable pre-filled cartridges (rechargeable)’

Sociodemographic characteristics included age (16-24/25-34/35-44/45-54/55-65/≥65 years), gender (men/women), occupational social grade (ABC1 includes managerial, professional, and upper supervisory occupations/C2DE includes manual routine, semi-routine, lower supervisory, and long-term unemployed), nation (England/Wales/Scotland), and history of ≥1 diagnosed mental health condition since the age of 16 (yes/no; assessed in waves up to June 2023 among all participants in England and ~50% of participants in Wales and Scotland).

Smoking status was assessed by asking participants which of the following best applied to them:

a. ‘I smoke cigarettes (including hand-rolled) every day’
b. ‘I smoke cigarettes (including hand-rolled), but not every day’
c. ‘I do not smoke cigarettes at all, but I do smoke tobacco of some kind (e.g., pipe, cigar or shisha)’
d. ‘I have stopped smoking completely in the last year’
e. ‘I stopped smoking completely more than a year ago’
f. ‘I have never been a smoker (i.e., smoked for a year or more)’

Those who responded *a* to *c* were considered current smokers, those who responded *d* recent (<1y) ex-smokers, *e* long-term (≥1 year) ex-smokers, and *f* never smokers.

Among current and recent ex-smokers (‘past-year smokers’), level of cigarette dependence was assessed with self-reported ratings of strength of urges to smoke over the past 24 hours [not at all (coded 0), slight (1), moderate (2), strong (3), very strong (4) and extremely strong (5)]. This variable was also coded 0 for smokers who responded ‘not at all’ to the (separate) question: ‘How much of the time have you spent with the urge to smoke?’.^24^

### Statistical analysis

Analyses were conducted using R v.4.2.1. They were not pre-registered and should be considered exploratory.

Survey weights were applied to match the sample to the demographic profile of Great Britain,^20,21^ with specific weights used for analyses of mental health conditions to account for this variable not being assessed among all participants in Wales and Scotland.

We excluded participants who did not respond to the questions on nicotine strength (those who responded that they did not know were included); those with missing data on other variables were excluded on a per-analysis basis (see figure legends and tables for information on sample sizes for each analysis).

#### Trend analyses

Using data from vapers aged ≥18 years in England surveyed between July 2016 and January 2024, we used logistic regression to test associations between survey wave and each response option (dummy coded) for nicotine strength of e-liquids used, including don’t know responses. Survey wave was modelled using restricted cubic splines with five knots, to allow relationships with time to be flexible and non-linear.

To explore moderation of trends by the main device type used, age, and smoking status, we repeated the models including the interaction between the moderator of interest and survey wave – thus allowing time trends to differ across sub-groups. Each interaction was tested in a separate model.

We used predicted estimates from these models to plot the proportions (with 95% confidence intervals [CI]) of vapers using each nicotine strength over time. On these figures, we included a vertical line indicating the timing of the rise in popularity of disposable e-cigarettes among young adults (estimated to be June 2021, based on previous findings^2,25^) to contextualise changes in the nicotine strengths being used.

#### Analyses of current nicotine strengths used

Using data from vapers aged ≥16 years in Great Britain surveyed between January 2022 and January 2024, we reported the proportions (with 95% CI) of vapers who reported using each different nicotine strength or who did not know, overall and by the main device type used, sociodemographic characteristics, smoking status, and (among past-year smokers) strength of urges to smoke.

We also calculated these separately stratified by the main device type used, to check whether the pattern of results differed between those mainly using refillable, disposable, and pod devices (we excluded strength of urges to smoke from these analyses, due to low numbers within subgroups).

Finally, we used multinomial logistic regression to test associations (among users of all device types) between nicotine strength and each participant characteristic in turn, adjusting for survey wave. For this analysis, we collapsed nicotine strengths to no nicotine, ≤11 mg/ml nicotine, and ≥12 mg/ml nicotine, to roughly approximate the proposed structure of the Vaping Products Duty.^10^ This was intended to offer insight into which groups would be most affected by the proposed duty.

## Results

A total of 9,286 vapers were surveyed in eligible waves, of whom 8,641 provided data on the nicotine strength of the e-liquid they mainly used. For the trend analyses, we selected those aged ≥18 and living in England, providing a sample of 7,314 participants (weighted mean [SD] age = 40.8 [15.2] years; 44.8% women; 54.0% social grades C2DE). For analyses of current nicotine strengths used in 2022-24, we selected those aged ≥16 years and living in Great Britain surveyed between January 2022 and January 2024, providing a sample of 2,373 participants (weighted mean [SD] age = 37.4 [15.3] years; 47.3% women; 54.1% social grades C2DE). In total, we analysed data from 7,957 unique participants.

### Trends in nicotine strength of e-liquids used by vapers in England

Figure 1 shows modelled trends in nicotine strengths of e-liquids used by vapers aged ≥18 years in England between July 2016 and January 2024. Figures 2, 3, and 4 show trends by the main device type used, age, and smoking status, respectively.

**Figure 1.**
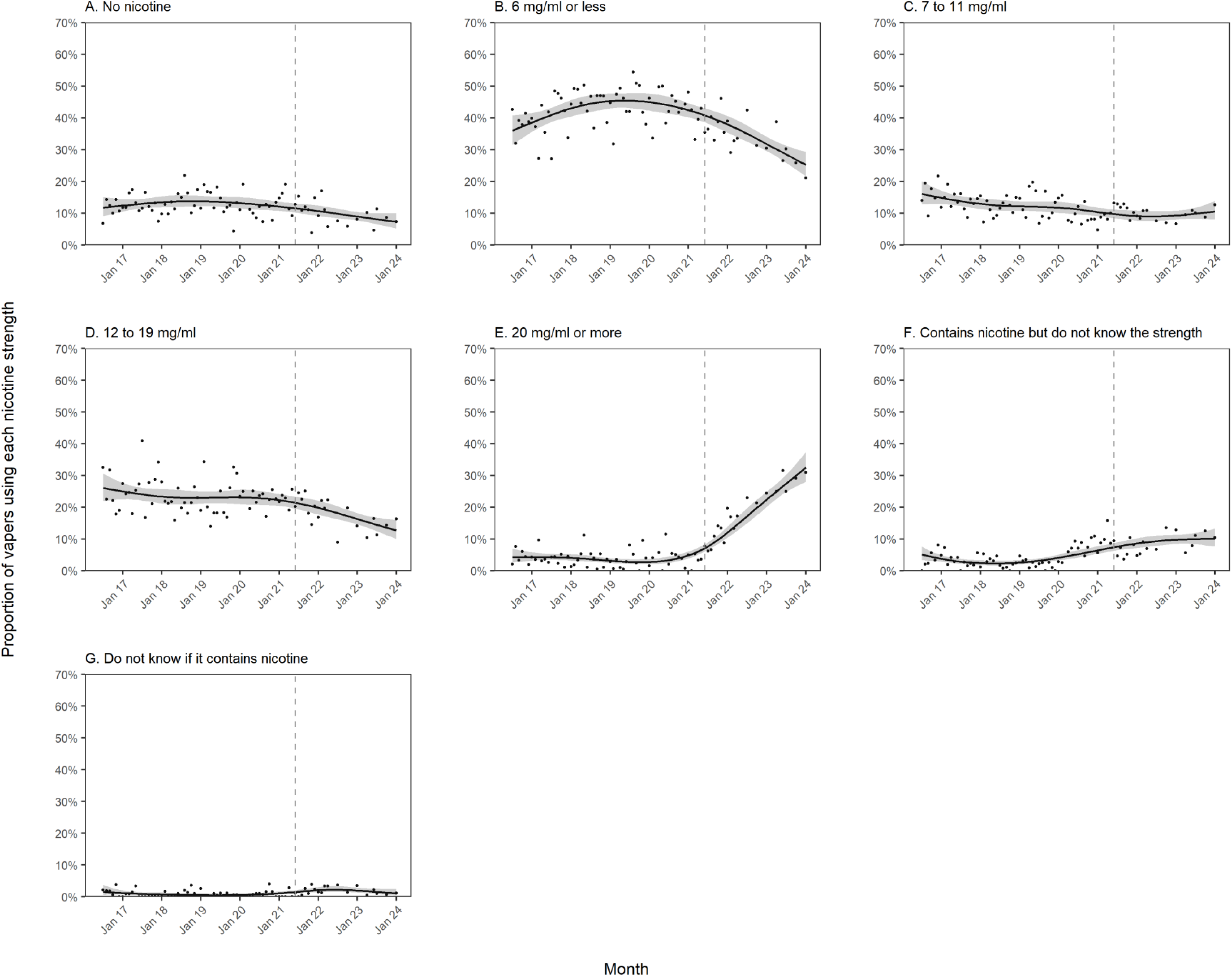
Trends in nicotine strengths of e-liquids used by adult (≥18y) vapers in England, July 2016 to January 2024. Unweighted sample size: *n*=7,314. Lines represent the modelled weighted proportion by monthly survey wave (modelled non-linearly using restricted cubic splines with five knots). Shaded bands represent 95% confidence intervals. Points represent the unmodelled weighted proportion by month. The vertical dashed line indicates the timing of the start of the rise in popularity of disposable vaping in June 2021.

**Figure 2.**
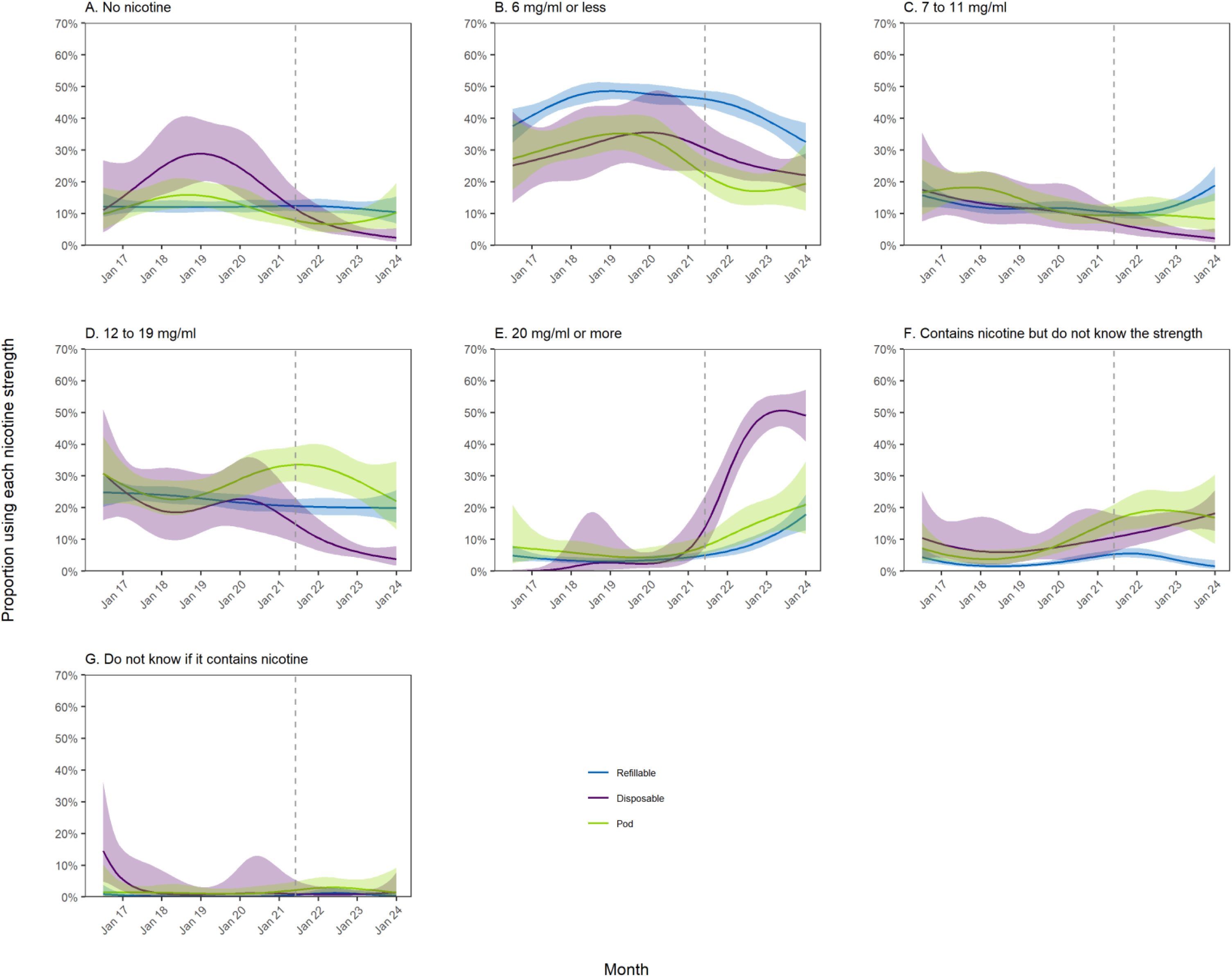
Trends in nicotine strengths of e-liquids used by adult (≥18y) vapers in England, July 2016 to January 2024, by the main device type used. Unweighted sample sizes: *n*=5,197 refillable; *n*=880 disposable; *n*=1,131 pod. Lines represent the modelled weighted proportion by monthly survey wave (modelled non-linearly using restricted cubic splines with five knots) and the main device type used. Shaded bands represent 95% confidence intervals. Points represent the unmodelled weighted proportion by month. The vertical dashed line indicates the timing of the start of the rise in popularity of disposable vaping in June 2021.

**Figure 3.**
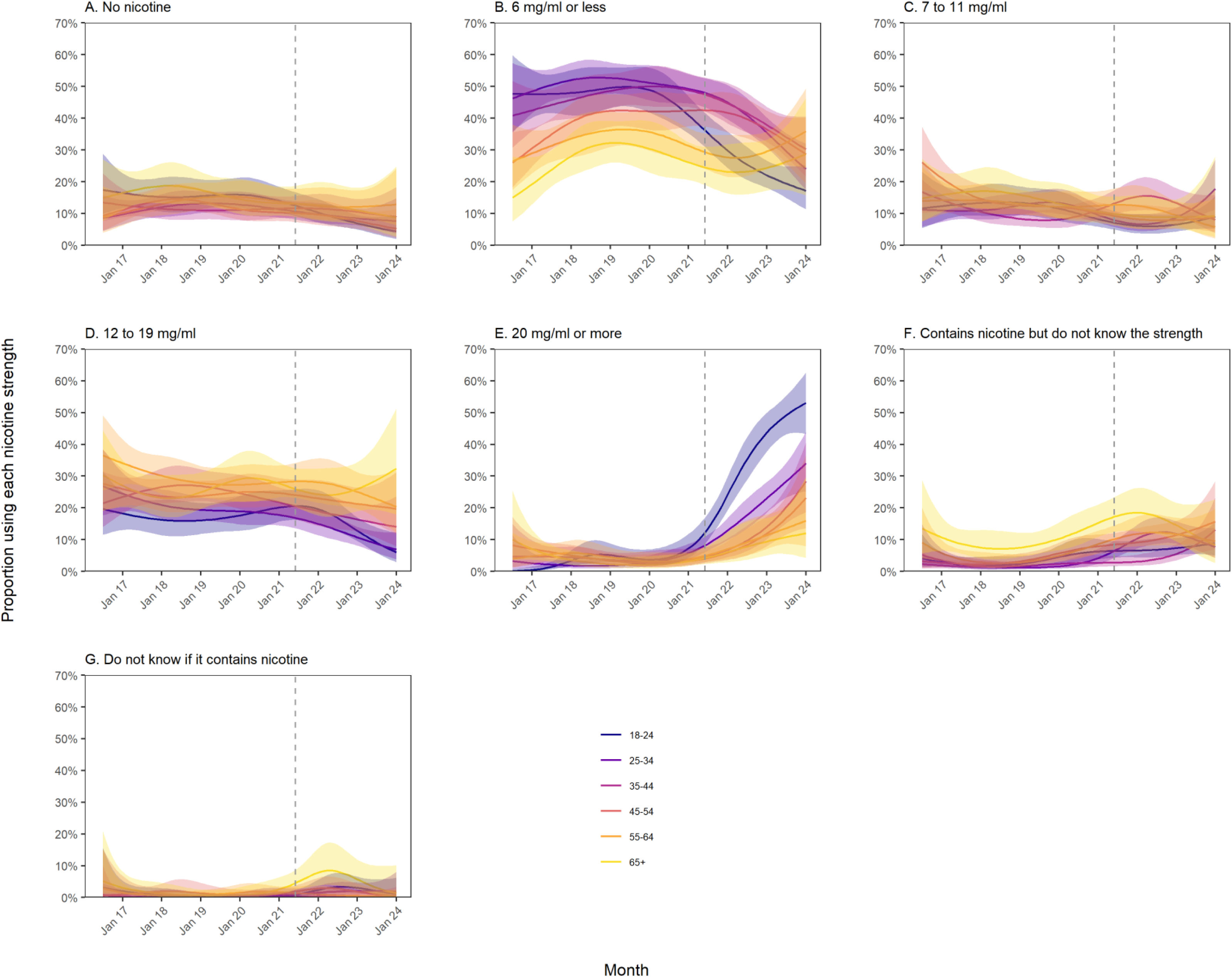
Trends in nicotine strengths of e-liquids used by adult (≥18y) vapers in England, July 2016 to January 2024, by age. Unweighted sample sizes: *n*=1,209 18-24-year-olds; *n*=1,650 25-34-year-olds; *n*=1,302 35-44-year-olds; *n*=1,337 45-54-year-olds; *n*=1,080 55-64-year-olds; *n*=736 ≥65-year-olds. Lines represent the modelled weighted proportion by monthly survey wave (modelled non-linearly using restricted cubic splines with five knots) and age. Shaded bands represent 95% confidence intervals. Points represent the unmodelled weighted proportion by month. The vertical dashed line indicates the timing of the start of the rise in popularity of disposable vaping in June 2021.

**Figure 4.**
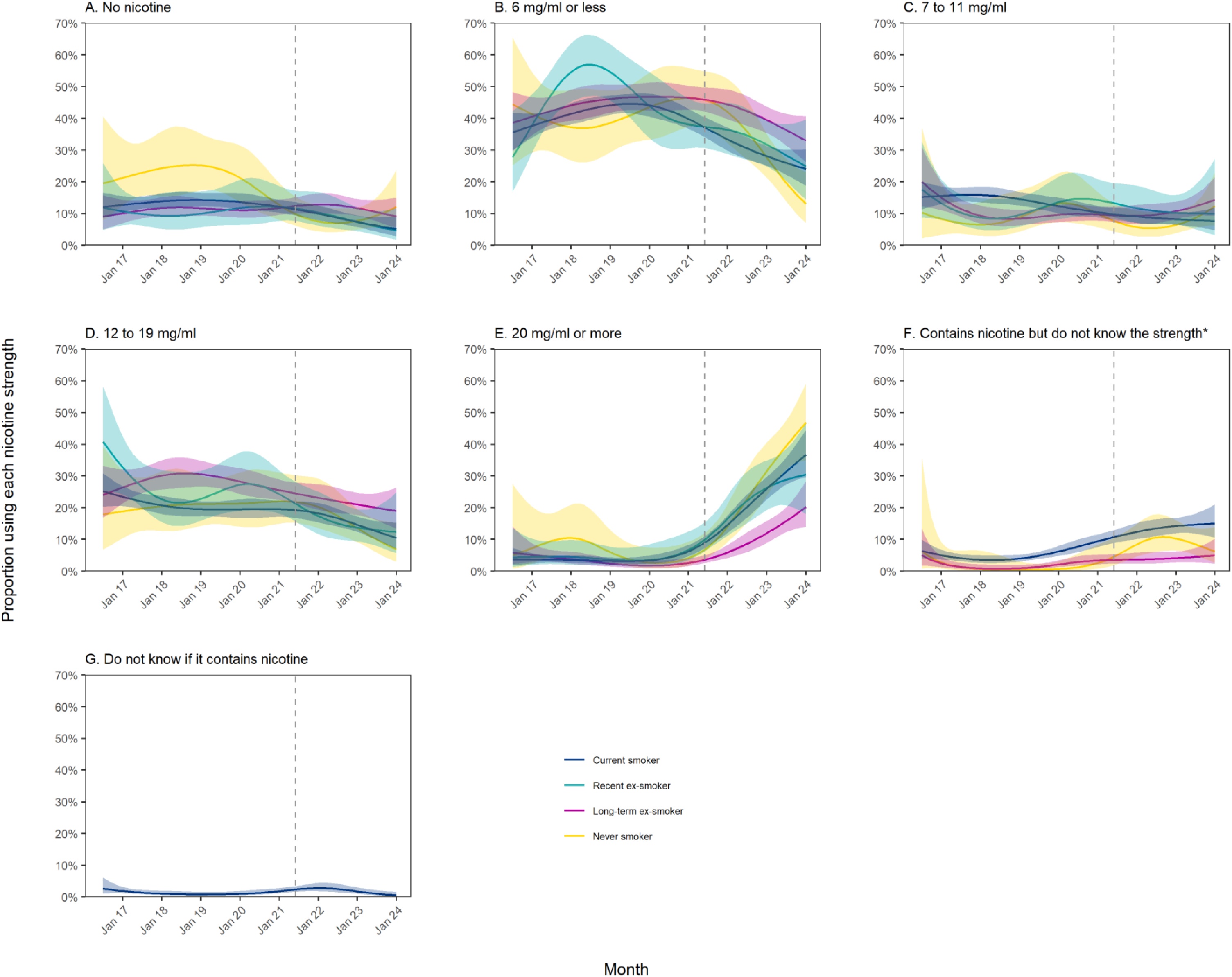
Trends in nicotine strengths of e-liquids used by adult (≥18y) vapers in England, July 2016 to January 2024, by smoking status. Unweighted sample sizes: *n*=530 never smokers; *n*=2,251 long-term (≥1y) ex-smokers; *n*=624 recent (<1y) ex-smokers; *n*=3,909 current smokers. Lines represent the modelled weighted proportion by monthly survey wave (modelled non-linearly using restricted cubic splines with five knots) and smoking status. Shaded bands represent 95% confidence intervals. Points represent the unmodelled weighted proportion by month. The vertical dashed line indicates the timing of the start of the rise in popularity of disposable vaping in June 2021. * Trends are not reported for recent ex-smokers in panel F or for never, long-term ex-, or recent ex-smokers in panel G because fewer than 30 participants in these groups endorsed these response options over the entire period, introducing substantial imprecision into the estimates.

The proportion of vapers who reported using the highest-strength (≥20 mg/ml) e-liquids increased from an average of 3.8% [95%CI 2.9–5.0%] up to June 2021 to 32.5% [27.9–37.4%] in January 2024 (Figure 1E). This was offset by declines over the same period in the proportion using nicotine-free (from 11.5% [10.2–13.0%] to 7.3% [5.2–10.0%]; Figure 1A), ≤6 mg/ml (from 40.8% [38.6–43.0%] to 25.3% [21.6–29.3%]; Figure 1B) and 12–19 mg/ml e-liquids (from 21.4% [19.6–23.4%] to 12.7% [10.0–16.0%]; Figure 1D), while the proportion using 7–11 mg/ml e-liquids remained relatively stable (at an average of 9.4% [7.9–11.3%]; Figure 1C).

The proportion who said that the e-cigarette they mainly used contained nicotine but they did not know the strength increased in 2020 and 2021, from an average of 3.1% [2.4–4.1%] up to December 2019 to 8.5% [7.2–10.0%] by December 2021, then remained relatively stable at an average of 9.6% [8.1–11.5%] between January 2022 and January 2024 (Figure 1F). The proportion who did not know if the e-cigarette they mainly used contained nicotine was low (<2.5%) across the period (Figure 1G).

The overall increase in the proportion using ≥20 mg/ml e-liquids since 2021 was observed across all device types but was greatest among those mainly using disposables (increasing from an average of 2.6% [0.9–9.3%] up to June 2021 to 49.0% [40.8–57.2%] in January 2024, compared with 3.7% [2.8–5.0%] to 17.9% [12.9–24.3%] and 5.7% [3.3–9.8%] to 21.0% [11.8–24.7%] among refillable and pod users, respectively (Figure 2E).

It was also more pronounced among vapers aged 18–24 than those in older age groups, increasing from an average of 3.9% [2.1–7.0%] up to June 2021 to 53.1% [43.3–62.7%] in January 2024, compared with 3.9% [2.0–7.6%] to 28.4% [18.7–40.7%] among those aged 35–44 and 3.8% [1.9–7.5%] to 12.1% [4.2–29.8%] among those aged ≥65 (age groups selected as examples; Figure 3E). The decline in the use of ≤6 mg/ml e-liquids was also greater in the youngest age group, falling from a high of 49.9% [43.8–56.0%] in June 2019 to 17.2% [11.3–25.2%] in January 2024, compared with 49.5% [44.1–54.8%] to 30.2% [21.6–40.5%] among those aged 35–44 and 32.0% [25.6–39.2%] to 28.8% [15.8–46.5%] among those aged ≥65 over the same period (Figure 3B).

While the increase in the proportion using ≥20 mg/ml e-liquids since 2021 was observed across all smoking statuses, it was smallest among long-term ex-smokers: it increased from an average of 3.2% [1.9–5.5%] up to June 2021 to 20.3% [14.0–28.3%] in January 2024, compared with 6.1% [2.6-14.6%] to 46.9% [34.9–59.2%] among never smokers, 4.5% [2.1–9.8%] to 30.5% [18.0–46.7%] among recent ex-smokers, and 4.0% [2.8–5.6%] to 36.8% [29.6– 44.5%] among current smokers (Figure 4E). The decline in the use of ≤6 mg/ml e-liquids was greatest among never smokers, falling from 45.8% [37.1–54.9%] in June 2021 to 13.1% [7.1–23.0%] in January 2024, compared with 45.9% [42.1–49.7%] to 33.0% [26.2–40.7%] among long-term ex-smokers, 37.4% [30.6–44.7%] to 24.9% [14.4–39.6%] among recent ex-smokers, and 37.2% [34.1–40.4%] to 24.1% [18.8–30.3%] among current smokers (Figure 4B).

The overall increase in the proportion who said they did not know the strength of their nicotine-containing e-cigarette was observed across all smoking statuses, rising from 0.4% [0.1–3.1%], 1.0% [0.4–2.0%], and 4.0% [3.1–5.2%] in January 2019 to 6.3% [2.7–13.9%], 5.0% [2.4–10.3%], and 15.1% [10.6–21.0%] in January 2024 among never, long-term ex-, and current smokers, respectively (Figure 4F). However, the increase was limited to those who mainly used pod and disposable devices (reaching 16.9% [8.6–30.5%] and 18.3% [12.7–25.5%], respectively, by January 2024), with little overall change among those who mainly used refillables (1.5% [0.7–3.5%]; Figure 2F).

### Nicotine strength of e-liquids currently used by vapers in Great Britain

**Table 1** provides data on the nicotine strength of e-liquids used by vapers aged ≥16 years in Great Britain in 2022-24, overall and in relation to participant characteristics.

**Table 1.**
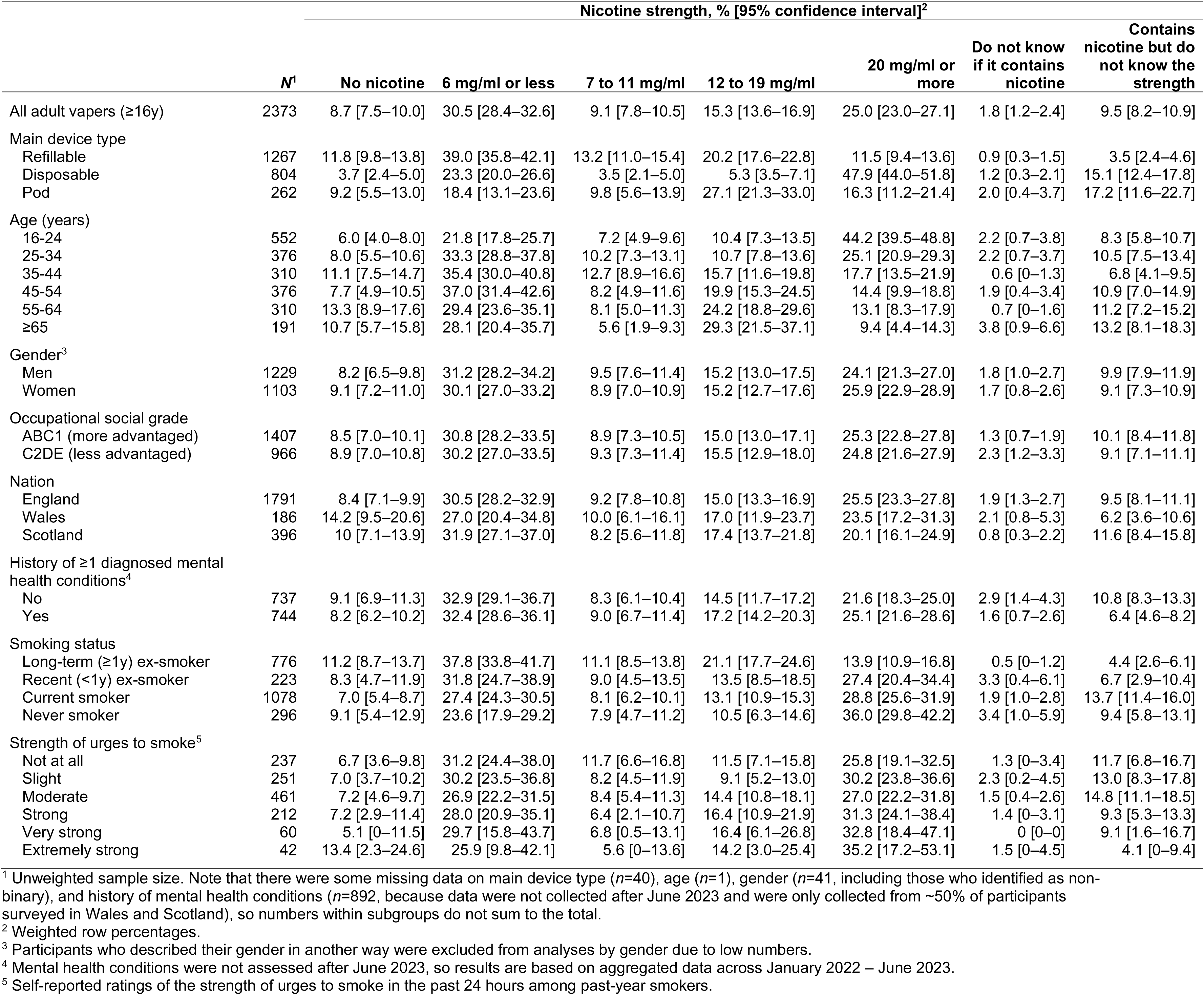
Usual nicotine strength used by adult (≥16y) vapers in Great Britain, January 2022-January 2024.

|  | N<sup>1</sup> | Nicotine strength, % [95% confidence interval]<sup>2</sup> |  |  |  |  |  | Contains nicotine but do not know the strength |
| --- | --- | --- | --- | --- | --- | --- | --- | --- |
|  |  | No nicotine | 6 mg/ml or less | 7 to 11 mg/ml | 12 to 19 mg/ml | 20 mg/ml or more | Do not know if it contains nicotine |  |
| All adult vapers (≥16y) | 2373 | 8.7 [7.5–10.0] | 30.5 [28.4–32.6] | 9.1 [7.8–10.5] | 15.3 [13.6–16.9] | 25.0 [23.0–27.1] | 1.8 [1.2–2.4] | 9.5 [8.2–10.9] |
| Main device type |  |  |  |  |  |  |  |  |
| Refillable | 1267 | 11.8 [9.8–13.8] | 39.0 [35.8–42.1] | 13.2 [11.0–15.4] | 20.2 [17.6–22.8] | 11.5 [9.4–13.6] | 0.9 [0.3–1.5] | 3.5 [2.4–4.6] |
| Disposable | 804 | 3.7 [2.4–5.0] | 23.3 [20.0–26.6] | 3.5 [2.1–5.0] | 5.3 [3.5–7.1] | 47.9 [44.0–51.8] | 1.2 [0.3–2.1] | 15.1 [12.4–17.8] |
| Pod | 262 | 9.2 [5.5–13.0] | 18.4 [13.1–23.6] | 9.8 [5.6–13.9] | 27.1 [21.3–33.0] | 16.3 [11.2–21.4] | 2.0 [0.4–3.7] | 17.2 [11.6–22.7] |
| Age (years) |  |  |  |  |  |  |  |  |
| 16-24 | 552 | 6.0 [4.0–8.0] | 21.8 [17.8–25.7] | 7.2 [4.9–9.6] | 10.4 [7.3–13.5] | 44.2 [39.5–48.8] | 2.2 [0.7–3.8] | 8.3 [5.8–10.7] |
| 25-34 | 376 | 8.0 [5.5–10.6] | 33.3 [28.8–37.8] | 10.2 [7.3–13.1] | 10.7 [7.8–13.6] | 25.1 [20.9–29.3] | 2.2 [0.7–3.7] | 10.5 [7.5–13.4] |
| 35-44 | 310 | 11.1 [7.5–14.7] | 35.4 [30.0–40.8] | 12.7 [8.9–16.6] | 15.7 [11.6–19.8] | 17.7 [13.5–21.9] | 0.6 [0–1.3] | 6.8 [4.1–9.5] |
| 45-54 | 376 | 7.7 [4.9–10.5] | 37.0 [31.4–42.6] | 8.2 [4.9–11.6] | 19.9 [15.3–24.5] | 14.4 [9.9–18.8] | 1.9 [0.4–3.4] | 10.9 [7.0–14.9] |
| 55-64 | 310 | 13.3 [8.9–17.6] | 29.4 [23.6–35.1] | 8.1 [5.0–11.3] | 24.2 [18.8–29.6] | 13.1 [8.3–17.9] | 0.7 [0–1.6] | 11.2 [7.2–15.2] |
| ≥65 | 191 | 10.7 [5.7–15.8] | 28.1 [20.4–35.7] | 5.6 [1.9–9.3] | 29.3 [21.5–37.1] | 9.4 [4.4–14.3] | 3.8 [0.9–6.6] | 13.2 [8.1–18.3] |
| Gender<sup>3</sup> |  |  |  |  |  |  |  |  |
| Men | 1229 | 8.2 [6.5–9.8] | 31.2 [28.2–34.2] | 9.5 [7.6–11.4] | 15.2 [13.0–17.5] | 24.1 [21.3–27.0] | 1.8 [1.0–2.7] | 9.9 [7.9–11.9] |
| Women | 1103 | 9.1 [7.2–11.0] | 30.1 [27.0–33.2] | 8.9 [7.0–10.9] | 15.2 [12.7–17.6] | 25.9 [22.9–28.9] | 1.7 [0.8–2.6] | 9.1 [7.3–10.9] |
| Occupational social grade |  |  |  |  |  |  |  |  |
| ABC1 (more advantaged) | 1407 | 8.5 [7.0–10.1] | 30.8 [28.2–33.5] | 8.9 [7.3–10.5] | 15.0 [13.0–17.1] | 25.3 [22.8–27.8] | 1.3 [0.7–1.9] | 10.1 [8.4–11.8] |
| C2DE (less advantaged) | 966 | 8.9 [7.0–10.8] | 30.2 [27.0–33.5] | 9.3 [7.3–11.4] | 15.5 [12.9–18.0] | 24.8 [21.6–27.9] | 2.3 [1.2–3.3] | 9.1 [7.1–11.1] |
| Nation |  |  |  |  |  |  |  |  |
| England | 1791 | 8.4 [7.1–9.9] | 30.5 [28.2–32.9] | 9.2 [7.8–10.8] | 15.0 [13.3–16.9] | 25.5 [23.3–27.8] | 1.9 [1.3–2.7] | 9.5 [8.1–11.1] |
| Wales | 186 | 14.2 [9.5–20.6] | 27.0 [20.4–34.8] | 10.0 [6.1–16.1] | 17.0 [11.9–23.7] | 23.5 [17.2–31.3] | 2.1 [0.8–5.3] | 6.2 [3.6–10.6] |
| Scotland | 396 | 10 [7.1–13.9] | 31.9 [27.1–37.0] | 8.2 [5.6–11.8] | 17.4 [13.7–21.8] | 20.1 [16.1–24.9] | 0.8 [0.3–2.2] | 11.6 [8.4–15.8] |
| History of ≥1 diagnosed mental health conditions<sup>4</sup> |  |  |  |  |  |  |  |  |
| No | 737 | 9.1 [6.9–11.3] | 32.9 [29.1–36.7] | 8.3 [6.1–10.4] | 14.5 [11.7–17.2] | 21.6 [18.3–25.0] | 2.9 [1.4–4.3] | 10.8 [8.3–13.3] |
| Yes | 744 | 8.2 [6.2–10.2] | 32.4 [28.6–36.1] | 9.0 [6.7–11.4] | 17.2 [14.2–20.3] | 25.1 [21.6–28.6] | 1.6 [0.7–2.6] | 6.4 [4.6–8.2] |
| Smoking status |  |  |  |  |  |  |  |  |
| Long-term (≥1y) ex-smoker | 776 | 11.2 [8.7–13.7] | 37.8 [33.8–41.7] | 11.1 [8.5–13.8] | 21.1 [17.7–24.6] | 13.9 [10.9–16.8] | 0.5 [0–1.2] | 4.4 [2.6–6.1] |
| Recent (<1y) ex-smoker | 223 | 8.3 [4.7–11.9] | 31.8 [24.7–38.9] | 9.0 [4.5–13.5] | 13.5 [8.5–18.5] | 27.4 [20.4–34.4] | 3.3 [0.4–6.1] | 6.7 [2.9–10.4] |
| Current smoker | 1078 | 7.0 [5.4–8.7] | 27.4 [24.3–30.5] | 8.1 [6.2–10.1] | 13.1 [10.9–15.3] | 28.8 [25.6–31.9] | 1.9 [1.0–2.8] | 13.7 [11.4–16.0] |
| Never smoker | 296 | 9.1 [5.4–12.9] | 23.6 [17.9–29.2] | 7.9 [4.7–11.2] | 10.5 [6.3–14.6] | 36.0 [29.8–42.2] | 3.4 [1.0–5.9] | 9.4 [5.8–13.1] |
| Strength of urges to smoke<sup>5</sup> |  |  |  |  |  |  |  |  |
| Not at all | 237 | 6.7 [3.6–9.8] | 31.2 [24.4–38.0] | 11.7 [6.6–16.8] | 11.5 [7.1–15.8] | 25.8 [19.1–32.5] | 1.3 [0–3.4] | 11.7 [6.8–16.7] |
| Slight | 251 | 7.0 [3.7–10.2] | 30.2 [23.5–36.8] | 8.2 [4.5–11.9] | 9.1 [5.2–13.0] | 30.2 [23.8–36.6] | 2.3 [0.2–4.5] | 13.0 [8.3–17.8] |
| Moderate | 461 | 7.2 [4.6–9.7] | 26.9 [22.2–31.5] | 8.4 [5.4–11.3] | 14.4 [10.8–18.1] | 27.0 [22.2–31.8] | 1.5 [0.4–2.6] | 14.8 [11.1–18.5] |
| Strong | 212 | 7.2 [2.9–11.4] | 28.0 [20.9–35.1] | 6.4 [2.1–10.7] | 16.4 [10.9–21.9] | 31.3 [24.1–38.4] | 1.4 [0–3.1] | 9.3 [5.3–13.3] |
| Very strong | 60 | 5.1 [0–11.5] | 29.7 [15.8–43.7] | 6.8 [0.5–13.1] | 16.4 [6.1–26.8] | 32.8 [18.4–47.1] | 0 [0–0] | 9.1 [1.6–16.7] |
| Extremely strong | 42 | 13.4 [2.3–24.6] | 25.9 [9.8–42.1] | 5.6 [0–13.6] | 14.2 [3.0–25.4] | 35.2 [17.2–53.1] | 1.5 [0–4.5] | 4.1 [0–9.4] |

<sup>1</sup> Unweighted sample size. Note that there were some missing data on main device type ( $n=40$ ), age ( $n=1$ ), gender ( $n=41$ , including those who identified as non-binary), and history of mental health conditions ( $n=892$ , because data were not collected after June 2023 and were only collected from ~50% of participants surveyed in Wales and Scotland), so numbers within subgroups do not sum to the total.
<sup>2</sup> Weighted row percentages.
<sup>3</sup> Participants who described their gender in another way were excluded from analyses by gender due to low numbers.
<sup>4</sup> Mental health conditions were not assessed after June 2023, so results are based on aggregated data across January 2022 – June 2023.
<sup>5</sup> Self-reported ratings of the strength of urges to smoke in the past 24 hours among past-year smokers.

Overall, 89.5% [95%CI 88.1–90.8%] said they usually used e-cigarettes containing nicotine, 8.7% [7.5–10.0%] said they used nicotine-free cigarettes, and 1.8% [1.2–2.4%] were unsure. The most commonly used nicotine strength was ≤6 mg/ml (30.5% of vapers), followed by ≥20 mg/ml (25.0%), 12–19 mg/ml (15.3%) and 7–11 mg/ml (9.1%).

However, the nicotine strength of e-liquids used differed across subgroups of vapers. There were clear differences between those using different e-cigarette device types. Almost half (47.9%) of disposable users reported using ≥20 mg/ml e-liquids compared with 16.3% of pod users and 11.5% of refillable users. By contrast, 50.8% of refillable users reported using ≤6 mg/ml or nicotine-free e-liquids, compared with 27.0% of disposable users and 27.6% of pod users. Refillable users were least likely to say that they did not know the strength of their nicotine-containing e-cigarette (3.5% vs. 15.1% and 17.2% of disposable and pod users, respectively).

There were also differences by age, with almost half (44.2%) of 16–24-year-olds using the highest strength (≥20 mg/ml) e-liquids, compared with 9.4–25.1% among older age groups. This age difference was observed across users of refillable (**Table S1**), disposable (**Table S2**), and pod (**Table S3**) devices.

Finally, there were differences by smoking status. Never smokers (36.0%), current smokers (28.8%), and recent (<1y) ex-smokers (27.4%) were more likely than long-term ex-smokers (13.9%) to report using the highest strength (≥20 mg/ml) e-liquids. They were also much more likely to say that they did not know whether their device contained nicotine (3.4%, 1.9%, and 3.3% vs. 0.5%, respectively) or that they did not know the strength of their nicotine-containing e-cigarette (9.4%, 13.7%, and 6.7% vs. 4.4%, respectively). The majority (87.4%) of never smokers were using nicotine-containing e-cigarettes, most commonly with ≥20 mg/ml e-liquids (36.0%). Of those using higher-strength e-liquids (≥12 mg/ml), never, recent ex-, and current smokers were more likely to be using ≥20 mg/ml than 12–19 mg/ml, but the opposite was true for long-term ex-smokers. A higher proportion of long-term ex-smokers than current and never smokers reported using low-nicotine (≤6 mg/ml) e-liquids (37.8% vs. 27.4% and 23.6%, respectively).

There were no notable differences in nicotine strength by gender, occupational social grade, history of mental health conditions, or (among smokers) level of cigarette dependence.

### Groups that would currently be most affected by the proposed Vaping Products Duty structure

**Table 2** shows the results of the multinomial logistic regression models, which summarise subgroup differences in the use of e-liquids that would be taxed at lower, intermediate, and higher levels according to the proposed Vaping Products Duty structure.

**Table 2.** Associations between usual nicotine strength and user characteristics among adult (≥16y) vapers in Great Britain, January 2022-January 2024.

|  | Nicotine strength vs. no nicotine,<br>OR [95% CI]<sup>1</sup> |  |
| --- | --- | --- |
|  | 11 mg/ml or less | 12 mg/ml or more |
| Main device type |  |  |
| Refillable | Ref | Ref |
| Disposable | 1.62 [1.10–2.38] | 5.21 [3.57–7.60] |
| Pod | 0.69 [0.44–1.09] | 1.75 [1.13–2.73] |
| Age (years) |  |  |
| 16-24 | Ref | Ref |
| 25-34 | 1.12 [0.73–1.72] | 0.49 [0.32–0.75] |
| 35-44 | 0.90 [0.58–1.39] | 0.33 [0.21–0.52] |
| 45-54 | 1.21 [0.73–2.02] | 0.49 [0.29–0.81] |
| 55-64 | 0.58 [0.36–0.96] | 0.31 [0.19–0.50] |
| $\geq 65$ | 0.65 [0.34–1.23] | 0.39 [0.21–0.73] |
| Gender<sup>2</sup> |  |  |
| Men | Ref | Ref |
| Women | 0.86 [0.65–1.14] | 0.94 [0.71–1.24] |
| Occupational social grade |  |  |
| ABC1 (more advantaged) | Ref | Ref |
| C2DE (less advantaged) | 0.96 [0.72–1.26] | 0.96 [0.72–1.27] |
| Occupational social grade |  |  |
| England | Ref | Ref |
| Wales | 0.55 [0.30–1.03] | 0.61 [0.33–1.13] |
| Scotland | 0.85 [0.51–1.41] | 0.79 [0.47–1.33] |
| History of $\geq 1$ diagnosed mental health conditions<sup>3</sup> | | |
| No | Ref | Ref |
| Yes | 1.10 [0.77–1.59] | 1.28 [0.89–1.85] |
| Smoking status |  |  |
| Long-term ($\geq 1$y) ex-smoker | Ref | Ref |
| Recent ($< 1$y) ex-smoker | 1.13 [0.68–1.87] | 1.61 [0.96–2.67] |
| Current smoker | 1.15 [0.84–1.58] | 1.93 [1.40–2.66] |
| Never smoker | 0.79 [0.51–1.22] | 1.59 [1.04–2.45] |
| Strength of urges to smoke<sup>4</sup> |  |  |
| Not at all | Ref | Ref |
| Slight | 0.86 [0.44–1.69] | 1.02 [0.52–2.02] |
| Moderate | 0.77 [0.43–1.39] | 1.06 [0.59–1.91] |
| Strong | 0.75 [0.37–1.51] | 1.20 [0.60–2.42] |
| Very strong | 1.12 [0.34–3.75] | 1.78 [0.54–5.89] |
| Extremely strong | 0.36 [0.12–1.06] | 0.64 [0.23–1.79] |

CI, confidence interval. OR, odds ratio.
<sup>1</sup> Odds ratio calculated using multinomial logistic regression (with no nicotine as the reference category), adjusted for survey wave.
<sup>2</sup> Participants who described their gender in another way were excluded from analyses by gender due to low numbers.
<sup>3</sup> Mental health conditions were not collected after June 2023, so results are based on data collected between January 2022 and June 2023.
<sup>4</sup> Self-reported ratings of the strength of urges to smoke in the past 24 hours among past-year smokers.

There were significant differences by the main device type used, age, and smoking status. Relative to those who mainly used refillable devices, disposable users had 1.62 times higher odds of using mid-strength (≤11 mg/ml) e-liquids and 5.21 times higher odds of using high-strength (≥12 mg/ml) e-liquids than nicotine-free e-liquids, and pod users had 1.75 times higher odds of using high-strength e-liquids.

The odds of using high-strength e-liquids vs. nicotine-free e-liquids were lower among older (≥25y) age groups than those aged 16-24y (OR range: 0.31-0.49) but the odds of using mid-strength e-liquids were similar across age groups.

Relative to long-term ex-smokers, current smokers had 93% higher odds of using high-strength vs. nicotine-free e-liquids, recent ex-smokers had 61% higher odds, and never smokers had 59% higher odds. The odds of using mid-strength e-liquids were more similar across smoking statuses.

There were no notable differences across other subgroups.

## Discussion

This study provides a comprehensive picture of the nicotine strengths of e-liquids used by adult vapers in Great Britain, with four key findings.

First, while nine in ten vapers reported using e-cigarettes that contain nicotine, there have been notable changes in the strengths used in England since 2016. In particular, there has been a sharp rise in the proportion of vapers using the highest-strength (≥20 mg/ml) e-liquids since disposable e-cigarettes started to become popular in the spring of 2021,^2,26^ offset by a decline in the proportion using lower-strength e-liquids (particularly ≤6 mg/ml). For the majority of the time the question was assessed, the question did not distinguish between 20 mg/ml (the legal limit) and higher concentrations. In the most recent data, the question distinguished between the two, and indicated that more than 90% of this group use the legal limit rather than stronger concentrations. While the increase in use of high-strength nicotine e-liquids was particularly pronounced among those using disposables, it was also observed across users of refillable and pod devices. It was greatest among 18–24-year-olds, consistent with the rise in use of disposable e-cigarettes being greatest at younger ages,^2,26^ and was similarly pronounced in people of all smoking statuses (including never smokers), except long-term ex-smokers.

Second, there has been an increase since 2020 in the proportion of vapers using disposable and pod devices who did not know how strong their nicotine-containing e-liquid was. It is possible that this increase is due to changes in where people are buying their vaping products since the Covid-19 pandemic. Before the pandemic, most vapers said that they usually bought their e-cigarettes and e-liquids from specialist vape shops,^27^ where staff are knowledgeable about the products and offer advice on nicotine strength.^28^ However, vape shops were forced to close during periods of lockdown,^29^ which saw a shift towards online purchasing.^27^ In addition, since disposable e-cigarettes were introduced to the market in 2021, supermarkets and convenience stores have become the most popular source of purchase.^27^ Typically, devices in these locations are stored behind a counter, and people cannot easily browse or inspect products before stating which device they would like to purchase. Better labelling and display by nicotine strength may be required to make the nicotine strength of products sold in these outlets clearer to consumers. If people are using illegal products, they may not clearly display nicotine content.

Third, the nicotine strength of e-liquids used currently tended to be higher among those who mainly use disposable devices and those aged 16–24 years (whether or not they are using disposables), and in people of all smoking statuses (including never smokers) except long-term ex-smokers. This suggests that the proposed Vaping Products Duty would disproportionately affect young vapers who have never smoked and may therefore contribute to reducing uptake (i.e., progression to regular use from experimentation) in this group, a stated policy objective. However, our results suggest it is not just never smokers who would be affected, but also current and recent ex-smokers who also tend to use higher nicotine strengths compared with long-term ex-smokers. This could have a number of unintended consequences.

If the duty discourages smokers from trying to quit with e-cigarettes or prompts them to use lower-strength e-liquids, it could undermine quitting and perpetuate smoking. Comparisons of the effectiveness of different doses of nicotine in e-cigarettes are limited.^13^ One randomised controlled trial (conducted in the USA where higher nicotine strengths are permitted than in the UK) found quit rates were 2.5 times higher among smokers randomised to receive an e-cigarette containing 36 mg/ml e-liquid than those who received 8 mg/ml, but the 95%CI included no difference (RR=2.50 [95%CI 0.80–7.77]).^14^ We found that while current smokers tended to use higher-strength e-liquids than long-term ex-smokers, one in three reported using low-strength (<6 mg/ml) or nicotine-free e-cigarettes. Given higher-strength e-liquids are more effective in relieving cravings for tobacco,^12^ people who want to use e-cigarettes to quit smoking could be encouraged to use higher-strength products (at least in the short term) to potentially increase their chances of quitting.^14^ However, the structure of the proposed duty will make it more expensive for smokers who do so. Our data do not tell us about the nicotine strength of e-liquids used by smokers in quit attempts, which may be higher than the average among current smokers and recent ex-smokers (as those who quit may reduce the nicotine strength used gradually after quitting).

If the duty encourages ex-smokers who vape to stop vaping or to switch to lower-strength e-liquids, there is a risk it could trigger relapse to smoking (although there is little direct evidence of the impact of e-cigarettes on long-term relapse, and people have also speculated that long-term nicotine dependence may be a greater risk factor for long-term relapse). This seems unlikely for long-term ex-smokers who reported using the lowest nicotine strengths, which may reflect people ‘tapering down’ their nicotine use over time or having quit with and continued using older-generation refillable devices (which, as we found, are typically used with lower-strength e-liquids than modern disposables). If it does not affect the risk of long-term relapse to smoking, then there are likely to be health benefits because vaping long-term is not harmless.^16^ However, higher strengths may be important for recent ex-smokers (who tended to use these), who may benefit from using high-strength nicotine e-liquid in the early phases of switching to minimise the risk of relapse.^30^

There is also a risk that the duty could worsen misperceptions about the harms of vaping. Recent data show smokers’ perceptions of the relative harms of e-cigarettes compared with cigarettes are as bad as they have ever been, with more than half believing they are equally or more harmful.^31^ Many people misattribute the cause of smoking-related disease to nicotine.^32,33^ Applying higher duty rates to higher-strength nicotine products may have the unwanted effect of worsening or maintaining these misperceptions if people think the tax is because the harms of these products are comparable to smoking rather than to reduce youth use.

The fourth key finding was that nicotine strength preferences did not differ substantially according to markers of disadvantage (e.g., by occupational social grade or history of mental health conditions) or by level of cigarette dependence among vapers who smoked. While this provides some reassurance that levying higher rates of tax on higher-strength e-liquids may not disproportionately affect disadvantaged or more dependent smokers who vape, our data only reflect vapers’ *current* nicotine strength preferences. They do not offer insight into how vapers’ choice of nicotine strength may change when the duty is introduced.

If any of the potential responses outlined above are greater among disadvantaged groups, it could have a negative equity impact. E-cigarettes are an important intervention for reducing smoking-related inequalities, because they offer a less harmful way of using nicotine^16^ without the need to quit nicotine altogether, which can be appealing for people with difficult lives who are not ready to consider total nicotine abstinence. Vaping is also cheaper than smoking^34^ and price is a motivator for those on low incomes. The UK Government’s ‘Swap to Stop’ initiative to provide a million free e-cigarette starter packs (alongside behavioural support to quit) is focused on reducing inequalities.^35^ The Vaping Products Duty will need to be carefully communicated so as not to dissuade people who could benefit most from taking up the offer of a free e-cigarette starter pack from their local stop smoking service.

Further research is urgently needed to understand the extent to which these potential unintended consequences are likely to occur and how they can be mitigated.

Strengths of the study include the representative sample and up-to-date data on nicotine strength preferences. There were also limitations. Data were not collected in Wales and Scotland before October 2020, so our analyses of time trends were restricted to vapers in England. Sample sizes within some subgroups were relatively small, which meant estimates were imprecise (as indicated by wide 95% CIs) and there may be differences in nicotine strength preferences between groups that we did not detect. In addition, the 20 mg/ml nicotine strength classification included e-liquids at and exceeding the maximum legal limit in Great Britain,^16^ so we were unable to analyse trends in the use of nicotine strengths that exceed the legal limit.

In conclusion, use of high-strength nicotine e-liquids in England has increased sharply since disposable e-cigarettes have become popular. Most adult vapers in Great Britain use e-cigarettes that contain nicotine but different subgroups use different nicotine strengths with the strength tending to be higher among those who mainly use disposable devices, those aged 16-24 years, and lower among long-term ex-smokers. In applying higher rates of tax to higher-strength nicotine e-liquids, the proposed Vaping Products Duty may be effective in reducing progression from experimentation to regular use and dependence among young adults (and potentially youth, who were not assessed here), including those who have never smoked. There may, however, be implications arising from the proposed duty for smokers trying to quit by vaping, which need taking into account before finalising the tax structure. Monitoring the outcomes and any unintended consequences from the policy will be important.

## Data Availability

Data are available on Open Science Framework

https://osf.io/gp9jv/

## Declarations

### Ethics approval

Ethical approval for the STS was granted originally by the UCL Ethics Committee (ID 0498/001). The data are not collected by UCL and are anonymised when received by UCL.

