## Supplementary material for "Nicotine strength of e-liquids used by adult vapers in Great Britain: a population survey 2016 to 2024": Table S1

**Table S1.** Usual nicotine strength used by adult (≥16y) vapers in Great Britain, January 2022-January 2024 – refillable device users

| | | **Nicotine strength, % [95% CI][2]** | | | | | | |
|---|---|---|---|---|---|---|---|---|
| | ***N*[1]** | **No nicotine** | **6 mg/ml or less** | **7 to 11 mg/ml** | **12 to 19 mg/ml** | **20 mg/ml or more** | **Do not know if it contains nicotine** | **Contains nicotine but do not know the strength** |
| All adult vapers (≥16y) | 1267 | 11.8 [9.8–13.8] | 39 [35.8–42.1] | 13.2 [11.0–15.4] | 20.2 [17.6–22.8] | 11.5 [9.4–13.6] | 0.9 [0.3–1.5] | 3.5 [2.4–4.6] |
| Age (years) | | | | | | | | |
| 16-24 | 159 | 15.3 [9.3–21.2] | 25.6 [17.7–33.6] | 15.3 [9.1–21.4] | 20.2 [12.5–27.9] | 20.4 [13.3–27.4] | 1.6 [0–4.0] | 1.7 [0–4.0] |
| 25-34 | 284 | 10.9 [6.8–15.0] | 43.1 [36.4–49.7] | 15.1 [10.3–20.0] | 12.7 [8.2–17.2] | 13.5 [9.0–17.9] | 1.5 [0–3.1] | 3.2 [0.8–5.6] |
| 35-44 | 226 | 12.3 [7.2–17.3] | 43.6 [36.2–51.0] | 15.2 [9.9–20.6] | 19.2 [13.2–25.2] | 8.8 [4.6–13.1] | 0 [0–0] | 0.9 [0–1.9] |
| 45-54 | 253 | 7.7 [4.2–11.2] | 44.8 [37.6–52.0] | 10.4 [5.6–15.1] | 22.9 [16.8–29.0] | 7.5 [3.3–11.8] | 0.8 [0–1.9] | 5.9 [2.3–9.4] |
| 55-64 | 219 | 12.2 [7.3–17.1] | 35.6 [28.3–42.9] | 10.9 [6.5–15.3] | 27.6 [20.8–34.5] | 8.9 [4.0–13.8] | 0.5 [0–1.5] | 4.3 [1.7–6.9] |
| ≥65 | 125 | 15.0 [7.5–22.5] | 30.0 [20.5–39.6] | 7.4 [2.0–12.8] | 29.8 [20.1–39.6] | 8.8 [2.3–15.4] | 0.7 [0–2.0] | 8.2 [3.2–13.3] |
| Gender[3] | | | | | | | | |
| Men | 712 | 11.8 [9.1–14.4] | 39.0 [34.8–43.2] | 12.6 [9.7–15.4] | 19.1 [15.8–22.5] | 12.9 [9.9–15.9] | 1.0 [0–1.9] | 3.7 [2.1–5.3] |
| Women | 538 | 11.8 [8.7–15.0] | 39.2 [34.4–44.0] | 14.2 [10.7–17.7] | 21.3 [17.1–25.5] | 9.6 [6.7–12.5] | 0.6 [0–1.3] | 3.3 [1.8–4.7] |
| Occupational social grade | | | | | | | | |
| ABC1 (more advantaged) | 751 | 11.1 [8.7–13.6] | 40.2 [36.3–44.1] | 13.6 [10.8–16.3] | 19.5 [16.4–22.7] | 11.3 [8.7–13.9] | 0.9 [0.2–1.6] | 3.4 [1.9–4.9] |
| C2DE (less advantaged) | 516 | 12.3 [9.2–15.5] | 38.0 [33.2–42.8] | 12.9 [9.5–16.2] | 20.7 [16.7–24.7] | 11.7 [8.4–14.9] | 0.9 [0–1.8] | 3.5 [2.0–5.1] |
| Nation | | | | | | | | |
| England | 907 | 11.2 [9.1–13.6] | 39.2 [35.7–42.7] | 13.6 [11.3–16.3] | 19.9 [17.1–23.0] | 11.9 [9.7–14.5] | 1 [0.5–2] | 3.3 [2.2–4.7] |
| Wales | 117 | 18.8 [12.2–27.9] | 31.0 [22.5–41.0] | 11.2 [6.1–19.6] | 22.9 [15.7–32.3] | 10.6 [5.9–18.3] | 0 [0–0] | 5.4 [2.3–12.0] |
| Scotland | 243 | 14.6 [10.1–20.6] | 41.4 [34.8–48.3] | 9.8 [6.2–15.2] | 21.9 [16.7–28.2] | 7.6 [4.6–12.3] | 0 [0–0] | 4.8 [2.4–9.3] |
| History of ≥1 diagnosed mental health conditions[4] | | | | | | | | |
| No | 412 | 10.9 [7.6–14.1] | 42.3 [37.0–47.6] | 10.8 [7.6–14.0] | 19.9 [15.5–24.2] | 10.4 [7.0–13.8] | 1.3 [0.1–2.5] | 4.4 [2.3–6.4] |
| Yes | 389 | 11.8 [8.5–15.0] | 40.8 [35.3–46.3] | 11.7 [8.1–15.3] | 21.2 [16.5–25.9] | 10.0 [6.7–13.3] | 1.1 [0–2.1] | 3.5 [1.6–5.5] |
| Smoking status | | | | | | | | |
| Long-term (≥1y) ex-smoker | 587 | 12.0 [9.0–15.0] | 45.0 [40.3–49.8] | 12.9 [9.6–16.1] | 21.3 [17.3–25.3] | 6.5 [4.1–9.0] | 0.5 [0–1.4] | 1.7 [0.3–3.1] |
| Recent (<1y) ex-smoker | 108 | 12.2 [5.8–18.5] | 35.5 [24.9–46.1] | 16.2 [7.4–25.0] | 20.8 [11.9–29.7] | 11.9 [5.2–18.5] | 1.6 [0–3.8] | 1.9 [0–4.7] |
| Current smoker | 459 | 10.1 [7.1–13.2] | 34.1 [29.0–39.2] | 12.5 [9.0–16.1] | 18.9 [14.8–22.9] | 16.0 [11.8–20.1] | 1.2 [0–2.4] | 7.1 [4.7–9.6] |
| Never smoker | 113 | 16.0 [7.8–24.1] | 32.5 [22.1–42.9] | 14.4 [7.2–21.5] | 19.4 [10.7–28.1] | 17.2 [9.5–24.9] | 0.6 [0–1.8] | 0 [0–0] |

[1] Unweighted sample size.
[2] Weighted row percentages.
[3] Participants who described their gender in another way were excluded from analyses by gender due to low numbers.
[4] Mental health conditions were not collected after June 2023, so results are based on aggregated data across January 2022 – June 2023.

**Table S2.** Usual nicotine strength used by adult (≥16y) vapers in Great Britain, January 2022-January 2024 – disposable device users

| | | Nicotine strength, % [95% CI][2] | | | | | | |
|---|---|---|---|---|---|---|---|---|
| | *N*[1] | No nicotine | 6 mg/ml or less | 7 to 11 mg/ml | 12 to 19 mg/ml | 20 mg/ml or more | Do not know if it contains nicotine | Contains nicotine but do not know the strength |
| All adult vapers (≥16y) | 804 | 3.7 [2.4–5.0] | 23.3 [20.0–26.6] | 3.5 [2.1–5.0] | 5.3 [3.5–7.1] | 47.9 [44.0–51.8] | 1.2 [0.3–2.1] | 15.1 [12.4–17.8] |
| Age (years) | | | | | | | | |
| 16-24 | 341 | 1.2 [0.2–2.2] | 21.1 [16.0–26.1] | 3.1 [1.1–5.1] | 5.3 [2.4–8.1] | 58.1 [52.1–64.0] | 1.2 [0–2.5] | 10.1 [6.6–13.6] |
| 25-34 | 199 | 4.6 [1.6–7.6] | 25.7 [18.8–32.5] | 3.8 [0.7–6.9] | 4.5 [1.3–7.6] | 44.1 [36.1–52.0] | 1.4 [0–3.7] | 16.0 [10.4–21.6] |
| 35-44 | 112 | 6.7 [1.7–11.6] | 24.8 [15.3–34.3] | 5.3 [0.1–10.6] | 5.3 [0.6–10.0] | 43.0 [32.5–53.5] | 1.1 [0–2.8] | 13.8 [6.8–20.8] |
| 45-54 | 77 | 4.4 [0–8.8] | 24.6 [14.3–34.9] | 1.8 [0–4.5] | 7.5 [0.6–14.4] | 38.7 [25.7–51.8] | 0 [0–0] | 23.1 [11.6–34.6] |
| 55-64 | 50 | 11.4 [2.0–20.8] | 19.5 [7.9–31.1] | 3.3 [0–8.2] | 4.4 [0–10.5] | 25.9 [11.4–40.5] | 1.3 [0–4.0] | 34.1 [19.4–48.9] |
| ≥65 | 25 | 2.5 [0–7.6] | 35.3 [10.1–60.4] | 4.9 [0–14.6] | 10.3 [0–24.5] | 16.4 [0.1–32.7] | 3.3 [0–8.2] | 27.5 [8.4–46.5] |
| Gender[3] | | | | | | | | |
| Men | 372 | 2.3 [0.9–3.8] | 21.8 [17.1–26.5] | 5.1 [2.6–7.7] | 6.1 [3.3–9.0] | 46.6 [40.7–52.4] | 1.1 [0–2.2] | 16.9 [12.6–21.2] |
| Women | 416 | 4.9 [2.8–7.0] | 24.9 [20.1–29.8] | 2.1 [0.6–3.7] | 4.3 [2.1–6.5] | 49.0 [43.6–54.5] | 1.3 [0–2.7] | 13.3 [9.9–16.8] |
| Occupational social grade | | | | | | | | |
| ABC1 (more advantaged) | 475 | 4.6 [2.6–6.6] | 21.9 [17.9–25.9] | 2.7 [1.1–4.3] | 5.2 [2.9–7.5] | 48.0 [43.2–52.9] | 0.6 [0–1.3] | 17.0 [13.3–20.6] |
| C2DE (less advantaged) | 329 | 3.0 [1.3–4.8] | 24.5 [19.3–29.6] | 4.2 [1.9–6.5] | 5.4 [2.8–8.0] | 47.8 [41.8–53.8] | 1.7 [0.2–3.2] | 13.5 [9.6–17.4] |
| Nation | | | | | | | | |
| England | 636 | 3.6 [2.5–5.3] | 23.8 [20.4–27.6] | 3.4 [2.2–5.4] | 5.5 [3.9–7.8] | 47.7 [43.5–52.0] | 1.1 [0.5–2.6] | 14.7 [12.1–17.9] |
| Wales | 51 | 4.0 [0.9–16.4] | 16.3 [8.4–29.3] | 2.5 [0.6–10.5] | 4.7 [1.0–19.6] | 59.5 [44.1–73.1] | 4.8 [1.3–16.1] | 8.0 [3.4–17.7] |
| Scotland | 117 | 4.8 [2.0–11.1] | 19.1 [12.3–28.4] | 4.8 [2.0–11.4] | 2.5 [0.9–7.0] | 45.5 [35.9–55.5] | 0.6 [0.1–3.9] | 22.6 [15.1–32.4] |
| History of ≥1 diagnosed mental health conditions[4] | | | | | | | | |
| No | 219 | 5.1 [2.1–8.0] | 24.6 [18.2–31.1] | 5.1 [1.8–8.4] | 4.3 [1.4–7.3] | 44.0 [36.7–51.3] | 0.7 [0–1.8] | 16.1 [10.8–21.3] |
| Yes | 258 | 2.8 [0.9–4.7] | 24.2 [18.7–29.8] | 4.5 [1.7–7.3] | 6.0 [2.7–9.4] | 51.8 [45.1–58.4] | 2 [0.3–3.6] | 8.7 [5.0–12.3] |
| Smoking status | | | | | | | | |
| Long-term (≥1y) ex-smoker | 105 | 5.9 [1.3–10.4] | 19.7 [11.4–27.9] | 4.2 [0.3–8.2] | 7.5 [1.4–13.6] | 50.4 [39.5–61.3] | 0.3 [0–0.8] | 12.1 [5.1–19.0] |
| Recent (<1y) ex-smoker | 86 | 3.1 [0–6.3] | 26.9 [16.1–37.7] | 0 [0–0] | 2.1 [0–5.5] | 52.2 [39.7–64.7] | 2.7 [0–8.0] | 12.9 [4.8–21.1] |
| Current smoker | 464 | 3.5 [1.7–5.3] | 24.9 [20.3–29.5] | 4.2 [2.0–6.3] | 6.5 [4–9.1] | 44.0 [38.9–49.2] | 0.7 [0–1.4] | 16.2 [12.5–19.8] |
| Never smoker | 149 | 3.3 [0.6–5.9] | 18.8 [11.8–25.7] | 3.1 [0.4–5.8] | 1.9 [0–4.6] | 55.3 [46.4–64.1] | 2.5 [0–5.1] | 15.2 [8.8–21.5] |

[1] Unweighted sample size.
[2] Weighted row percentages.
[3] Participants who described their gender in another way were excluded from analyses by gender due to low numbers.
[4] Mental health conditions were not collected after June 2023, so results are based on aggregated data across January 2022 – June 2023.

**Table S3.** Usual nicotine strength used by adult (≥16y) vapers in Great Britain, January 2022-January 2024 – pod device users

| | *N*[1] | Nicotine strength, % [95% CI][2] | | | | | | |
|---|---|---|---|---|---|---|---|---|
| | | **No nicotine** | **6 mg/ml or less** | **7 to 11 mg/ml** | **12 to 19 mg/ml** | **20 mg/ml or more** | **Do not know if it contains nicotine** | **Contains nicotine but do not know the strength** |
| All adult vapers (≥16y) | 262 | 9.2 [6.1–13.7] | 18.4 [13.7–24.2] | 9.8 [6.3–14.8] | 27.1 [21.7–33.4] | 16.3 [11.8–22.0] | 2.0 [0.9–4.5] | 17.2 [12.3–23.4] |
| Age (years) | | | | | | | | |
| 16-24 | 46 | 7.6 [2.7–19.3] | 14.5 [6.6–29.0] | 10.0 [4.0–23.1] | 15.2 [6.8–30.5] | 33.1 [19.9–49.5] | 4.2 [1.0–16.2] | 15.4 [6.9–30.7] |
| 25-34 | 51 | 2.8 [0.7–10.9] | 15.3 [6.8–30.9] | 11.1 [4.6–24.5] | 24.3 [13.8–39.2] | 19.6 [10.1–34.6] | 1.6 [0.2–11.1] | 25.3 [13.6–42.1] |
| 35-44 | 51 | 10.4 [4.2–23.5] | 23.7 [13.4–38.4] | 18.1 [8.3–35.1] | 23.2 [13.0–37.8] | 8.8 [3.0.–22.7] | 0 [0–0] | 15.9 [6.9–32.5] |
| 45-54 | 37 | 14.6 [5.9–31.9] | 23.1 [11.0–42.3] | 7.9 [2.6–21.2] | 34.4 [20.1–52.2] | 7.4 [2.1–22.7] | 1.0 [0.1–7.7] | 11.6 [3.5–32.1] |
| 55-64 | 39 | 21.8 [9.7–42.0] | 14.7 [5.8–32.6] | 2.3 [0.3–15.8] | 38.7 [23.2–56.9] | 14.2 [5.7–31.1] | 0 [0–0] | 8.3 [3.1–20.4] |
| ≥65 | 38 | 4.4 [1.0–17.9] | 20.6 [9.3–39.7] | 1.3 [0.2–9.3] | 39.5 [23.0–58.7] | 7.5 [2.3–21.9] | 6.6 [1.9–20.3] | 20.1 [9.2–38.5] |
| Gender[3] | | | | | | | | |
| Men | 121 | 5.9 [2.9–11.6] | 22.5 [15.1–32.2] | 8.3 [4.1–16.0] | 26.6 [19.0–35.9] | 17.1 [10.6–26.5] | 2.6 [0.9–7.4] | 16.9 [10.0–27.2] |
| Women | 134 | 10.8 [6.1–18.3] | 14.7 [9.4–22.3] | 11.6 [6.6–19.6] | 28.3 [20.6–37.5] | 16.2 [10.5–24.3] | 0.9 [0.2–4.0] | 17.5 [11.2–26.3] |
| Occupational social grade | | | | | | | | |
| ABC1 (more advantaged) | 163 | 9.7 [5.8–15.9] | 19.9 [13.8–27.7] | 8.3 [4.9–13.8] | 26.3 [19.8–34.1] | 18.7 [12.7–26.6] | 1.8 [0.5–5.9] | 15.4 [10.4–22.1] |
| C2DE (less advantaged) | 99 | 8.8 [4.5–16.3] | 16.8 [10.2–26.3] | 11.3 [5.8–20.8] | 28.0 [19.5–38.4] | 13.8 [7.9–23.0] | 2.3 [0.8–6.5] | 19.0 [11.2–30.4] |
| Nation | | | | | | | | |
| England | 214 | 9.6 [6.3–14.4] | 17.9 [13–24.2] | 9.2 [5.6–14.6] | 26.3 [20.5–33.0] | 17.5 [12.6–23.6] | 1.9 [0.8–4.6] | 17.7 [12.5–24.4] |
| Wales | 16 | 14.3 [2.6–50.6] | 34.5 [10.9–69.4] | 25.6 [7.7–58.7] | 15.0 [3.8–44.1] | 3.7 [0.4–26.2] | 0 [0–0] | 6.9 [1.4–28.5] |
| Scotland | 32 | 0 [0–0] | 16.5 [6.4–36.6] | 11.4 [3.9–28.9] | 50.0 [31.3–68.7] | 2.9 [0.4–19.0] | 5.6 [1.3–21.0] | 13.7 [5.3–30.9] |
| History of ≥1 diagnosed mental health conditions[4] | | | | | | | | |
| No | 86 | 13.4 [7.6–22.5] | 19.3 [11.7–30.2] | 7.3 [2.9–17.1] | 20.9 [13.3–31.3] | 16.5 [9.6–26.8] | 2.8 [0.9–8.7] | 19.7 [11.2–32.4] |
| Yes | 86 | 4.6 [1.7–12.1] | 19.8 [12.3–30.3] | 12.1 [6.0–23.2] | 35.7 [25.7–47.2] | 14.2 [8.0–24.0] | 0.9 [0.1–6.5] | 12.6 [7.2–21.1] |
| Smoking status | | | | | | | | |
| Long-term (≥1y) ex-smoker | 80 | 12.8 [6.8–22.7] | 14.1 [7.8–24.2] | 9.6 [4.1–20.8] | 40.3 [28.9–53.0] | 10.9 [5.3–21.0] | 0.4 [0.1–3.2] | 11.8 [5.4–23.9] |
| Recent (<1y) ex-smoker | 25 | 7.5 [1.7–27.5] | 34.9 [16.4–59.3] | 14.2 [4.8–35.2] | 28.6 [13.3–51.2] | 5.2 [0.7–31.4] | 4.8 [0.6–29.3] | 4.7 [0.6–29.1] |
| Current smoker | 132 | 7.3 [3.6–14.1] | 18.6 [12.2–27.4] | 10.2 [5.4–18.3] | 20.5 [14.2–28.8] | 19.7 [13.1–28.6] | 1.5 [0.5–5] | 22.2 [14.8–31.9] |
| Never smoker | 25 | 10.9 [3.3–30.7] | 13.9 [4.9–33.7] | 3.9 [0.8–16.7] | 22.1 [8.9–45.4] | 24.4 [9.9–48.6] | 6.8 [1.5–25.7] | 18.0 [6.0–42.9] |

[1] Unweighted sample size.
[2] Weighted row percentages.
[3] Participants who described their gender in another way were excluded from analyses by gender due to low numbers.
[4] Mental health conditions were not collected after June 2023, so results are based on aggregated data across January 2022 – June 2023.
